# Development and evaluation of a live birth prediction model for evaluating human blastocysts: a retrospective study

**DOI:** 10.1101/2022.10.20.22281296

**Authors:** Hang Liu, Zhuoran Zhang, Yifan Gu, Changsheng Dai, Guanqiao Shan, Haocong Song, Daniel Li, Wenyuan Chen, Ge Lin, Yu Sun

**Affiliations:** Department of Mechanical Engineering, University of Toronto, 5 King’s College Road, Toronto, Ontario, M5S 3G8, Canada; Department of Electrical and Computer Engineering, 5 King’s College Road, Toronto, Ontario, M5S 3G8, Canada; Institute of Biomedical Engineering, University of Toronto, 5 King’s College Road, Toronto, Ontario, M5S 3G8, Canada; Department of Computer Science, University of Toronto, 5 King’s College Road, Toronto, Ontario, M5S 3G8, Canada; School of Science and Engineering, The Chinese University of Hong Kong-Shenzhen, 2001 Longxiang Boulevard, Longgang District, Shenzhen, 518172, China; Institute of Reproduction and Stem Cell Engineering, Central South University, Changsha, China; Reproductive and Genetic Hospital of Citic-Xiangya, Changsha, China; Key Laboratory of Stem Cells and Reproductive Engineering, Ministry of Health, Changsha, China; National Engineering and Research Center of Human Stem Cell, Changsha, China

## Abstract

**Background:** In infertility treatment, blastocyst morphological grading is commonly used in clinical practice for blastocyst evaluation and selection, but has shown limited predictive power on live birth outcomes of blastocysts. To improve live birth prediction, a number of artificial intelligence (AI) models have been established. Most existing AI models for blastocyst evaluation only used images for live birth prediction, and the area under the receiver operating characteristic (ROC) curve (AUC) achieved by these models has plateaued at ∼0.65.

**Methods:** This study proposed a multi-modal blastocyst evaluation method using both blastocyst images and patient couple’s clinical features (e.g., maternal age, hormone profiles, endometrium thickness, and semen quality) to predict live birth outcomes of human blastocysts. To utilize the multi-modal data, we developed a new AI model consisting of a convolutional neural network (CNN) to process blastocyst images and a multi-layer perceptron to process patient couple’s clinical features. The dataset used in this study consists of 17,580 blastocysts with known live birth outcomes, blastocyst images, and patient couple’s clinical features.

**Results:** This study achieved an AUC of 0.77 for live birth prediction, which significantly outperforms related works in the literature. Sixteen out of 103 clinical features were identified to be predictors of live birth outcomes and helped improve live birth prediction. Among these features, maternal age, the day of blastocyst transfer, antral follicle count, retrieved oocyte number, and endometrium thickness measured before transfer are the top five features contributing to live birth prediction. Heatmaps showed that the CNN in the AI model mainly focuses on image regions of inner cell mass and trophectoderm (TE) for live birth prediction, and the contribution of TE-related features was greater in the CNN trained with the inclusion of patient couple’s clinical features compared with the CNN trained with blastocyst images alone.

**Conclusions:** The results suggest that the inclusion of patient couple’s clinical features along with blastocyst images increases live birth prediction accuracy.

**Funding:** Natural Sciences and Engineering Research Council of Canada and the Canada Research Chairs Program.

## Introduction

Infertility is a global health issue, affecting more than 50 million couples worldwide (*Mascarenhas et al., 2012*). Since the birth of the first in vitro fertilization (IVF) child in 1978, over 8 million children were born with IVF treatment (*Adamson et al., 2011*). Among the various factors contributing to IVF outcomes, the quality of the blastocyst (day-5 embryo) selected for transfer is critical for the success of IVF treatment. Manual grading of blastocyst development stage, inner cell mass (ICM), and trophectoderm (TE) remains the most common method for blastocyst evaluation. While the blastocyst morphological grading is widely used in clinical practice, morphological grades of the development stage, ICM, and TE have shown limited predictive power on clinical outcomes (*Seli et al., 2011; Reignier et al., 2019; Bartolacci et al., 2021; Ueno et al., 2021; Xiong et al., 2022*). It is desired to identify features for accurate prediction of clinical outcomes of blastocysts.

To achieve this goal, the convolutional neural network (CNN) is expected to play a critical role. CNN is able to automatically detect discriminative features from images and has been the state-of-the-art method in various fields in medical imaging, such as lung cancer prediction (*Ardila et al., 2019*), breast cancer prediction (*McKinney et al., 2020*), and diabetic retinopathy screening (*Bora et al., 2021*). To apply CNN to predict the clinical outcome of a blastocyst, images of blastocysts with a known clinical outcome (e.g., pregnancy, live birth) are collected for the CNN model development. The area under the receiver operating characteristic (ROC) curve (AUC) is the most commonly used metric to evaluate and compare machine learning models on predicting clinical outcomes of blastocysts (*Kragh et al., 2021*). The AUCs reported in the literature using CNN to predict clinical outcomes from blastocyst images range from 0.64 to 0.71 for pregnancy prediction (*VerMilyea et al., 2020; Kragh et al., 2021; Berntsen et al., 2022; Enatsu et al., 2022; Loewke et al., 2022*), and are around 0.65 for live birth prediction (*Miyagi et al., 2019, Nagaya et al., 2021*).

Besides using blastocyst images, attempts have also been made to use time-lapse videos for live birth prediction. These videos contain the entire development process from day 0 to day 5-7. However, results in the literature show that CNN using static images achieved similar or slightly better accuracies in predicting clinical outcomes than using time-lapse videos, for instance, AUC= 0.68-0.71 (*VerMilyea et al., 2020; Enatsu et al., 2022; Loewke et al., 2022*) vs. 0.64-0.67 (*Kragh et al., 2021; Berntsen et al., 2022*) for pregnancy prediction, and 0.66 (*Miyagi et al., 2019*) vs. 0.65 (*Nagaya et al., 2021*) for live birth prediction. A potential reason is that the redundant frames of images in time-lapse videos may work as noise causing the model to overfit and thus leading to a lower prediction accuracy (*Zhu et al., 2018; Wu et al., 2021; Tao et al., 2022*). Therefore, we opted to use static blastocyst images in this study.

Different from using blastocyst images alone to predict clinical outcomes, Miyagi et al. proposed to use blastocyst images together with maternal clinical features including maternal age, AMH, and BMI and reported an AUC of 0.74, the highest accuracy in literature (*Miyagi et al., 2020*). However, two questions remain elusive. First, the contribution of blastocyst images and the additional contribution of maternal clinical features to live birth prediction are unknown. Second, endometrium status-related features, such as endometrium thickness and pattern are also critical factors impacting live birth outcomes (*Ng et al., 2007; Bu et al., 2016; Mahutte et al., 2022*), but were not considered.

In this study, we quantified the effect of blastocyst images and the combined effect of both blastocyst images and patient couple’s clinical features on live birth prediction. The live birth prediction model using only blastocyst images achieved an AUC of 0.67, which was significantly outperformed by the AUC of 0.77 achieved by the model using both blastocyst images and patient couple’s clinical features (p-value < 0.0001). Additionally, when endometrium status-related features (e.g., endometrium thickness and pattern) were excluded, the AUC of the model using both blastocyst images and patient couple’s clinical features significantly decreased to 0.74 (p-value < 0.0001), indicating that the inclusion of endometrium status-related clinical features helps improve live birth prediction accuracy. Sixteen patient couple’s clinical features were identified to be most related to live birth outcomes of blastocysts, among which maternal age, the day of blastocyst transfer, antral follicle count (AFC), retrieved oocyte number, and endometrium thickness measured before transfer are the top five features contributing to live birth prediction. Additionally, the CNN heatmaps showed that the CNN mainly focused on ICM and TE for live birth prediction, and the contribution of TE-related features was greater in the CNN trained with the inclusion of patient couple’s clinical features compared with the CNN trained with blastocyst images alone.

## Methods

### Dataset collection

We used retrospectively collected data to develop the live birth prediction model. Transferred blastocysts with known live birth outcomes for patients who underwent frozen embryo transfer (FET) cycles from 2016 to 2020, at the Reproductive and Genetic Hospital of CITIC-Xiangya, were reviewed for inclusion in the dataset. This study was approved by the Ethics Committee of the Reproductive and Genetic Hospital of CITIC-Xiangya (approval number: LL-SC-2021-008).

Blastocyst images were captured before transfer using a standard optical light microscope mounted with a camera. Two grayscale images were captured for each blastocyst, one focusing on ICM and the other focusing on TE. Blastocysts were cropped from the original images which have a resolution of 1024×768 and were consistently padded to 500×500 to facilitate model training. Patient couple’s clinical features consist of 103 features including maternal age and BMI, the day of blastocyst transfer, infertility diagnosis and treatment history of patient couples, ovarian stimulation protocols, maternal hormone profiles and ultrasound results measured during the ovarian stimulation process and before transfer, and paternal semen diagnosis results (see Table I in Supplementary file 1 for a complete list). Based on p-value analysis and sequential forward feature selection (*Solorio-Fernández et al., 2020*, see Supplementary file 1 for implementation details), 16 clinical features that are most relevant to live birth prediction were identified and used for training the machine learning model (see Figure 3).

**Table I.** P-value analysis result of 103 patient couple’s clinical features. Fifty-one out of the 103 clinical features, highlighted in bold text, were identified to have statistical significance between blastocysts with positive and negative live birth outcomes. Y: yes, N: No.

| ID | Feature name | P-value $\leq 0.05$ |
| --- | --- | --- |
| Categorical features |  |  |
| **1** | **Infertility type** | **Y** |
| 2 | Tubal blockage | N |
| **3** | **Ovulatory dysfunction** | **Y** |
| **4** | **Endometriosis** | **Y** |
| 5 | Oligozoospermia | N |
| 6 | Asthenospermia | N |
| 7 | Oligoasthenozoospermia | N |
| 8 | Teratozoospermia | N |
| 9 | Maternal immune infertility | N |
| 10 | Obstructive azoospermia | N |
| **11** | **Azoospermia** | **Y** |
| 12 | Abnormal acrosome | N |
| 13 | Paternal chromosome abnormalities | N |
| **14** | **Ovarian hyperstimulation syndrome** | **Y** |
| **15** | **Poor endometrial receptivity** | **Y** |
| 16 | Maternal single-gene genetic disease | N |
|  | Maternal blood type |  |
| 17 | A | N |
| 18 | AB | N |
| 19 | B | N |
| 20 | O | N |
|  | Paternal blood type |  |
| 21 | A | N |
| **22** | **AB** | **Y** |
| 23 | B | N |
| 24 | O | N |
|  | Endometrium (EM) pattern before transfer |  |
| **25** | **A** | **Y** |
| 26 | B | N |
| 27 | C | N |
|  | EM pattern on HCG day |  |
| **28** | **A** | **Y** |
| **29** | **B** | **Y** |
| **30** | **C** | **Y** |
|  | Ovarian stimulation (OS) protocols |  |
| 31 | GnRH agonist long protocol | N |
| 32 | Clomiphene citrate | N |
| 33 | Progesterone | N |
| **34** | **Mild stimulation** | **Y** |
| **35** | **GnRH antagonists** | **Y** |
| 36 | Letrozole | N |
| **37** | **Short effect long scheme** | **Y** |
| **38** | **Natural cycle** | **Y** |
| 39 | Dydrogesterone | N |
| 40 | Short down-regulated | N |
| 41 | Ultra-short down-regulated | N |
| **42** | **Long down-regulated** | **Y** |
| **43** | **Ultra-long down-regulated** | **Y** |
| 44 | Ultra-ultra-long down-regulated | N |
|  | Insemination method |  |
| **45** | **IVF** | **Y** |
| 46 | ICSI | N |
| 47 | Combined | N |
|  | Sperm source |  |
| **48** | **Frozen semen** | **Y** |
| **49** | **Fresh semen** | **Y** |
| **50** | **Testicular sperm aspiration** | **Y** |
| **51** | **Percutaneous epididymal sperm aspiration** | **Y** |
|  | Sperm preparation techniques |  |
| 52 | Swim up | N |
| 53 | Hypo-osmotic swelling | N |
| **54** | **Density gradient centrifugation** | **Y** |
| 55 | Wash | N |
| 56 | Wash and swim up | N |
| Numerical features |  |  |
| **57** | **Maternal age** | **Y** |
| **58** | **Maternal body mass index** | **Y** |
| **59** | **Infertility duration** | **Y** |
| **60** | **Number of OS cycles** | **Y** |
| **61** | **EM thickness before transfer** | **Y** |
| **62** | **Prolactin before transfer** | **Y** |
| **63** | **Progesterone before transfer** | **Y** |
| **64** | **Estradiol before transfer** | **Y** |

| ID | Feature name | P value $\leq 0.05$ |
| --- | --- | --- |
| Continued |  |  |
| 65 | Number of OS days | N |
| 66 | **Days from OS to HCG day** | **Y** |
| 67 | **Days from OS to ovum pick-up** | **Y** |
| 68 | **EM thickness on HCG day** | **Y** |
| 69 | **Prolactin before transfer** | **Y** |
| 70 | **Progesterone on HCG day** | **Y** |
| 71 | **Estradiol on HCG day** | **Y** |
| 72 | **Luteinising hormone on HCG day** | **Y** |
| 73 | **Left antral follicle count** | **Y** |
| 74 | **Right antral follicle count** | **Y** |
| 75 | **Total antral follicle count** | **Y** |
| 76 | **Follicle stimulating hormone on day 3 after period** | **Y** |
| 77 | **Luteinising hormone on day 3 after period** | **Y** |
| 78 | Estradiol on day 3 after period | N |
| 79 | Prolactin on day 3 after period | N |
| 80 | Progesterone on day 3 after period | N |
| 81 | Testosterone on day 3 after period | N |
| 82 | Free triiodothyronine on day 3 after period | N |
| 83 | **Free thyroxine on day 3 after period** | **Y** |
| 84 | **Anti-Müllerian hormone on day 3 after period** | **Y** |
| 85 | Cancer antigen 125 | N |
| 86 | Blood sugar | N |
| 87 | Thyroid-stimulating hormone on day 3 after period | N |
| 88 | **Ratio of progesterone to estradiol on HCG day** | **Y** |
| 89 | **Retrieved oocyte number** | **Y** |
| 90 | Days of abstinence | N |
| 91 | **Day of blastocyst transfer** | **Y** |
| 92 | **Semen volume before processing** | **Y** |
| 93 | Semen volume after processing | N |
| 94 | Sperm density before processing | N |
| 95 | Sperm density after processing | N |
| 96 | Ratio of grade A sperms before processing | N |
| 97 | **Ratio of grade A sperms after processing** | **Y** |
| 98 | Ratio of grade B sperms before processing | N |
| 99 | **Ratio of grade B sperms after processing** | **Y** |
| 100 | Ratio of grade C sperms before processing | N |
| 101 | Ratio of grade C sperms after processing | N |
| 102 | Ratio of grade D sperms before processing | N |
| 103 | Ratio of grade D sperms after processing | N |

A total of 28,118 blastocysts with known live birth outcomes were reviewed, among which 17,580 blastocysts with two blastocyst images and all the 16 clinical features available were included in the dataset.

### Model architecture

Figure 1 shows the architecture of our live birth prediction model based on multi-modal blastocyst evaluation. It consists of a CNN to process blastocyst images and an MLP to process patient couple’s clinical features. The decision-level feature from the CNN and the decision-level feature from the MLP each have two variables used to classify the blastocyst into the positive or negative live birth outcome category. The adding operation fuses decision-level features from the CNN and the MLP, and the addition result is taken as the final output of the overall live birth prediction model.

**Figure 1.**
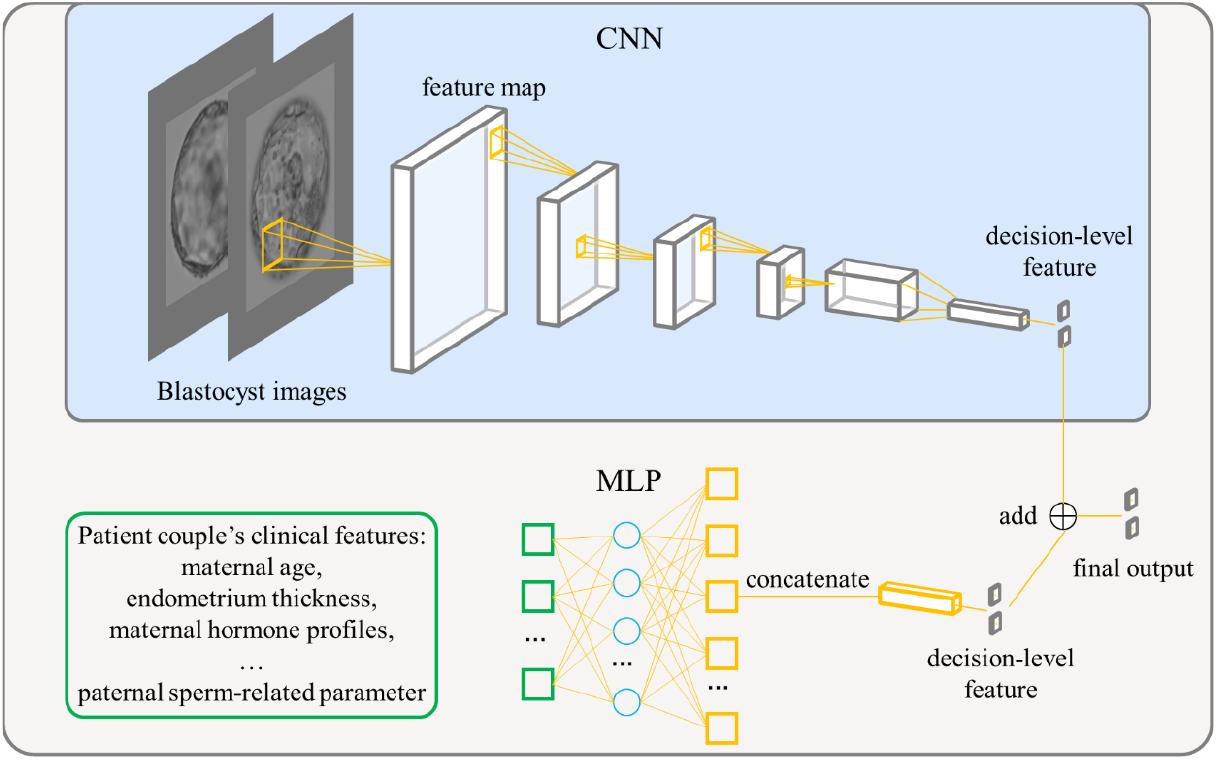
Architecture of our live birth prediction model based on multi-modal blastocyst evaluation. CNN: convolutional neural network; MLP: multi-layer perceptron.

### Model implementation and training

The proposed live birth prediction model used EfficientNetV2-S as the backbone CNN. EfficientNetV2-S is the baseline model in the EfficientNetV2 family, which is a new family of CNN models that provide higher accuracy and training speed than conventional models (*Tan et al., 2021*). In our work, the output dimension of the final fully connected layer in EfficientNetV2-S was set to be two, representing the positive and negative live birth outcome of a blastocyst, respectively. The model was implemented using PyTorch 1.10.1 (*Paszke et al., 2019*).

Each of the 17,580 blastocysts had two images taken at different focal planes, one focused more on TE cells and the other on ICM. Furthermore, for each blastocyst, live birth outcomes and all the 16 patient couple’s clinical features were available. The blastocysts were randomly split into 80%:10%:10% to construct the training, validation, and testing datasets. Since the ratio of blastocysts with a positive live birth outcome in the dataset is 0.368, to mitigate the model’s prediction bias towards the majority category (i.e., the negative live birth outcome), the weighted sampling approach, which can help rebalance the class distributions while sampling from an imbalanced dataset (*Feng et al., 2021*), was employed for training the model.

### Statistical analysis

Statistical tests were calculated to compare clinical features between blastocysts with the positive live birth outcome and blastocysts with the negative live birth outcome. Chi-squared test was used for categorial features, T-test was used for numerical features. Chi-squared test and T-test were performed using Python (Version: 3.6). ROC curves were compared by the DeLong test implemented in MedCalc software(Version: 20). All statistical tests were two-tailed and considered significant if p-value ≤ 0.05.

## Results

### The inclusion of patient couple’s clinical features increased AUC for live birth prediction

To quantify the individual effect of blastocyst images and the combined effect of patient couple’s clinical features, we built and compared models that 1) used only blastocyst images for live birth prediction, and 2) used both blastocyst images and patient couple’s clinical features for prediction. In addition, to quantify the specific effect of endometrium status-related features (i.e., endometrium thickness before transfer, endometrium thickness on HCG day, and endometrium pattern B (yes/no) on HCG day) on live birth prediction, a third model trained using blastocyst images and patient couple’s clinical features where endometrium status-related features were excluded, was also built and compared.

Figure 2 shows the ROC curves and AUCs of the three models for predicting live birth outcomes of 1,758 blastocysts (i.e., 10% of 17,580) in the test dataset. Using only blastocyst images for live birth prediction gave an AUC of 0.67, with a 95% confidence interval (CI) of 0.72 to 0.76. Using blastocyst images and patient couple’s clinical features (endometrium status-related features excluded) significantly increased the AUC to 0.74 (95% CI: 0.72-0.76, p-value < 0.0001). Using both blastocyst images and patient couple’s clinical features (endometrium status-related features included) achieved a prediction AUC of 0.77 (95% CI: 0.75-0.79), which is significantly higher than using only blastocyst images for prediction (p-value < 0.0001) and than using blastocyst images and patient couple’s clinical features where endometrium status-related features were excluded (p-value = 0.007).

**Figure 2.**
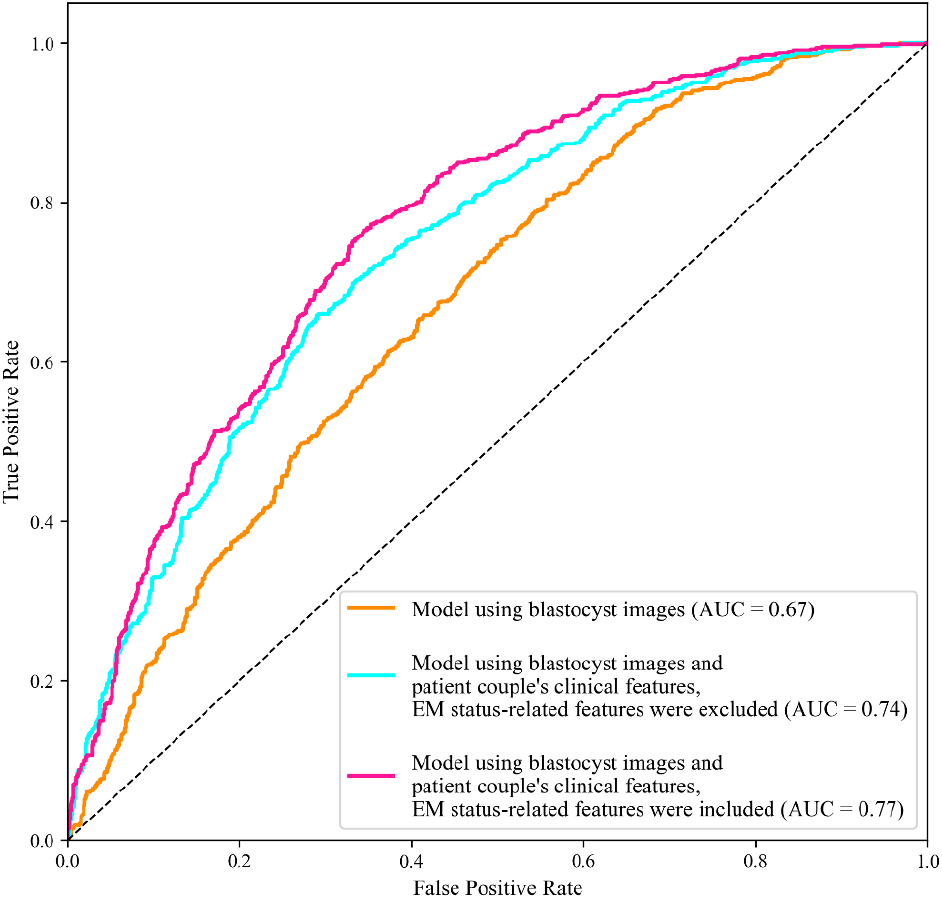
Receiver operating characteristic (ROC) analysis. ROC curves of the model using only blastocyst images, the model using blastocyst images and patient couple’s clinical features where EM-status related features were excluded, and the model using blastocyst images and patient couple’s clinical features where EM-status related features were included to predict live birth outcomes of 1,758 blastocysts in the test dataset. AUC: area under the ROC curve; EM: endometrium; EM status-related features: endometrium thickness before transfer, endometrium thickness on HCG day, endometrium pattern B (yes/no) on HCG day. **Source data 1**. Code and data used to generate the ROC curves.

### Ranking the predictive power of patient couple’s clinical features

We then investigated the predictive power of each patient couple’s clinical feature in predicting live birth outcome. Figure 3 shows the 16 features that were identified to be most related to the live birth outcomes of the blastocysts. These features were ranked according to their AUCs for individually predicting the live birth outcomes of blastocysts using univariable logistic regression. The AUC for each feature was reported as the mean AUC over a 10-fold cross-validation process.

**Figure 3.**
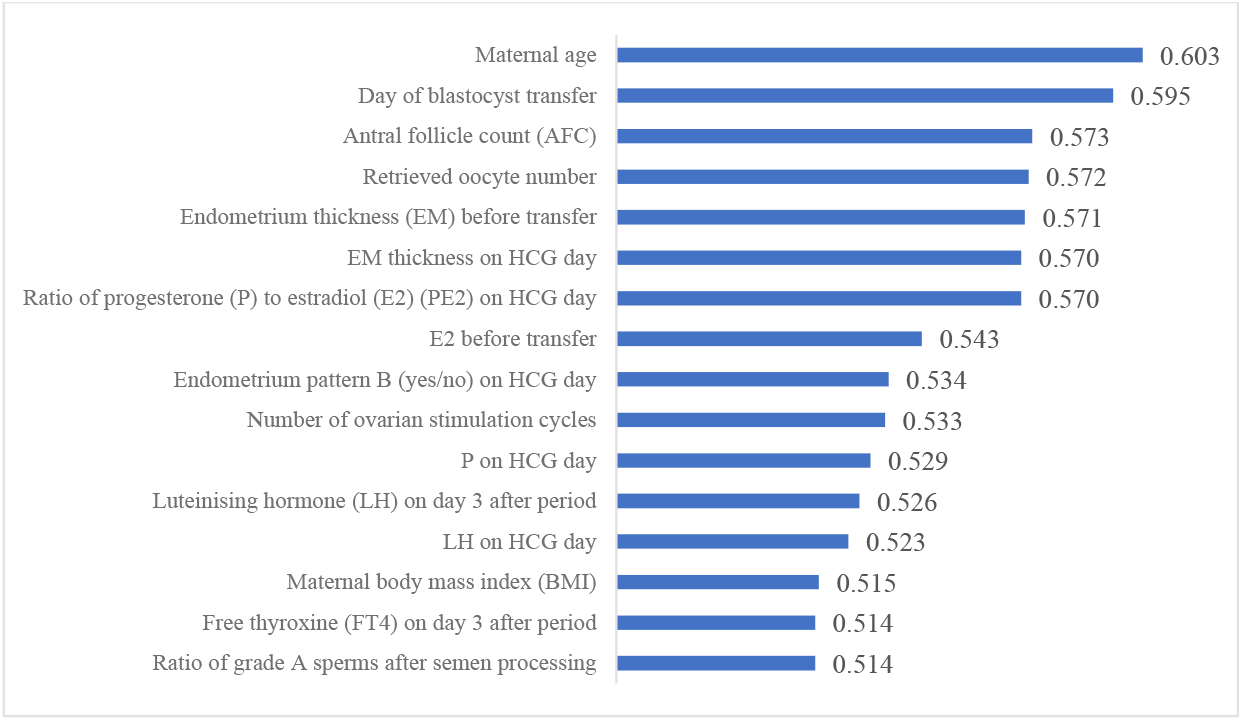
Ranking the predictive power of patient couple’s clinical features. The 16 patient couple’s clinical features that were identified to be most related to the live birth outcomes of the blastocysts ranked by the AUC for individually predicting live birth outcome. **Source data 1**. Source data to generate the AUC ranking chart.

### CNN heatmaps

In blastocyst images, what does CNN focus on to predict the live birth outcome of a blastocyst? Is there a difference in what the CNN focuses on between the model trained without and with the inclusion of patient couple’s clinical features? To answer these questions, we used the class activation mapping method to generate heatmaps. Figure

4 shows blastocyst images, corresponding heatmaps of the CNN trained without including patient couple’s clinical features, and corresponding heatmaps of the CNN trained with including patient couple’s clinical features.

The blastocyst images were cropped and padded to a consistent size to facilitate model training. The padding value was calculated as the mean pixel value of blastocyst images in the dataset. Heatmaps were generated by XGradCAM (*Fu et al., 2020*). Note that the CNN takes two-channel blastocyst images as the input, one focusing on ICM and the other focusing on TE. The blastocyst images shown in Figure 4 are those focused on ICM, and Supplemental Figure 1 shows the two-channel blastocyst images. As can be seen in Figure 4, when trained using only blastocyst images, the CNN mainly focuses on ICM and TE for predicting live birth outcomes. When training with both blastocyst images and patient couple’s clinical features, TE-related features contributed more to live birth prediction compared with training with blastocyst images only.

**Figure 4.**
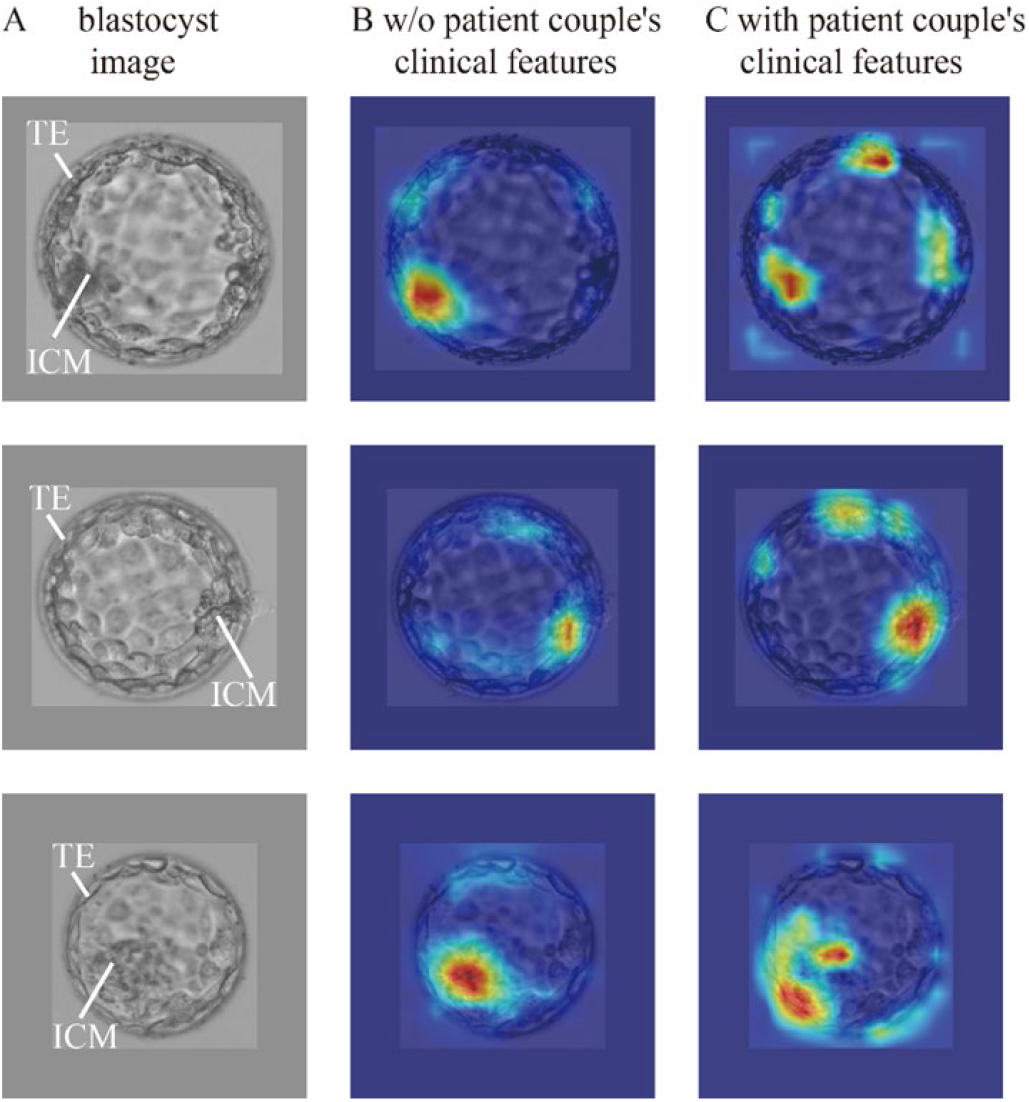
CNN heatmaps analysis. Heatmaps of the CNN trained without and with patient couple’s clinical features. Column **A**: original blastocyst images. Column **B**: corresponding heatmaps of the CNN trained without including patient couple’s clinical features. Column **C**: corresponding heatmaps of the CNN trained with the inclusion of patient couple’s clinical features. **Source code 1**. Code and data used to generate the heatmaps in columns B, C.

## Discussion

In this study, the individual effect of blastocyst images and the combined effect of patient couple’s clinical features for live birth prediction were quantified by comparing the AUC values of the model using only blastocyst images and the model using both blastocyst images and patient couple’s clinical features. An AUC of 0.67 was achieved with blastocyst images only while using both blastocyst images and patient couple’s clinical features led to a significantly higher AUC of 0.77 in live birth prediction. When endometrium status-related features were excluded from patient couple’s clinical features, the AUC of the live birth prediction model significantly decreased (p-value = 0.007) from 0.77 to 0.74, indicating the strong relevance of endometrium status-related features in live birth prediction. Sixteen patient couple’s clinical features were identified to be most related to live birth outcomes of blastocysts, among which maternal age, the day of blastocyst transfer, AFC, retrieved oocyte number, and endometrium thickness before transfer are the top five features contributing to live birth prediction.

This study was based on a comprehensive multi-modal dataset collected for blastocyst evaluation. The dataset includes 17,580 blastocysts with known live birth outcomes, blastocyst images, and 16 patient couple’s clinical features. As shown in Figure 3, the 16 patient couple’s clinical features comprehensively include maternal basal characteristics (age, BMI); hormone profiles measured after period (LH, FT4), on HCG day (PE2, P, LH), and before transfer (E2); endometrium status-related features (endometrium thickness on HCG day and before transfer, endometrium pattern on HCG day); features related to oocytes (AFC, retrieved oocyte number); the day of blastocyst transfer; number of ovarian stimulation cycles; and paternal features (the ratio of grade A sperm after semen processing). For comparison, the dataset studied by Miyagi et al. (*Miyagi et al., 2020*) did not contain endometrium status-related features and key hormone profiles (e.g., P, E2, LH). There are numerous IVF datasets containing over 100,000 records of clinical features and live birth outcomes (*Nelson et al., 2011; McLernon et al., 2016; La et al., 2021*); however, there are no blastocyst images in these datasets, and thus, these datasets cannot be used for building models to evaluate blastocysts from their images.

To handle the multi-modal dataset, our proposed model was designed to integrate two modules including a CNN and an MLP to enable the model to simultaneously consider images and numerical clinical features for blastocyst evaluation. The large and comprehensive multi-modal dataset and the proposed CNN+MLP model resulted in the highest AUC value of 0.77 ever reported thus far for predicting live birth outcomes of blastocysts. They also enabled us to quantify the predictive power of each feature in predicting the live birth outcomes of blastocysts.

The blastocyst grading system introduced in 1999 (*Gardner 1999, Gardner and Schoolcraft, 1999*) remains the most common method used by embryologists to evaluate blastocyst quality although the morphological grades of blastocyst development stage, ICM and TE have limited predictive power on live birth outcomes (e.g., AUC= 0.58-0.61 for live birth prediction reported by *Reignier et al., 2019; Bartolacci et al., 2021; Xiong et al., 2022*). Since CNN became a state-of-the-art method for image-based classification, many attempts have been made to apply the CNN to blastocyst evaluation for predicting clinical outcomes (e.g., *VerMilyea et al., 2020; Kragh et al., 2021; Berntsen et al., 2022; Enatsu et al., 2022; Loewke et al., 2022; Miyagi et al., 2019; Nagaya et al., 2021*). Among these, the AUC values reported in the literature using blastocyst images only were around 0.65 for live birth prediction (*Miyagi et al., 2019; Nagaya et al., 2021*). Similarly, we achieved an AUC of 0.67 (see Figure 2). Compared with the AUC of 0.58-0.61 reported in the literature using Gardner grades for live birth prediction, these results confirmed that CNN can achieve a higher prediction accuracy. As shown in Figure 4, the CNN mainly focuses on ICM and TE. Different from the Gardner-defined TE grade on the number of TE cells and the cohesiveness of TE cells as a whole, the CNN tends to focus on specific TE clusters. Understanding the heatmaps further requires more investigations.

Miyagi et al. used both blastocyst images and maternal clinical features (age, AMH, BMI) to predict live birth outcomes and achieved an AUC of 0.74 (*Miyagi et al., 2020*). The additional contribution from the three maternal clinical features was not clear since no AUC was reported by using blastocyst images alone. Furthermore, despite their importance in pregnancy and live birth, endometrium status-related features were not considered in their work. Therefore, our study used a comprehensive dataset and quantitatively compared the AUC values of live birth prediction using only blastocyst images versus using both blastocyst images and patient couple’s clinical features. We also quantified the usefulness of endometrium status-related features in working with blastocyst images to improve live birth prediction. Furthermore, we revealed that hormone profiles such as E2, LH, P, and FT4, features related to oocyte retrieval such as AFC and number of oocytes retrieved, and the ratio of grade A sperm after processing representing semen quality are able to work with blastocyst images to further improve the live birth prediction accuracy.

Another finding of this study is that the weights of TE-related features increased by comparing the heatmaps of the CNN trained without and with including patient couple’s clinical features (see Figure 4). A potential reason may be that TE and the endometrium status-related features (e.g., endometrium thickness and pattern) play critical roles when a blastocyst starts the process of implantation, and a positive live birth outcome is not possible without the success of this implantation process (*Ahlström et al., 2011; Hill et al., 2013; Chen et al., 2014; Bakkensen et al., 2019*).

In conclusion, in this retrospective study involving 17,580 blastocysts with known live birth outcomes, blastocyst images and 16 patient couple’s clinical features, we built a live birth prediction model based on multi-modal blastocyst evaluation using both blastocyst images and patient couple’s clinical features. We quantified the individual effect of blastocyst images and the combined effect of patient couple’s clinical features on live birth prediction. Results demonstrated that using both blastocyst images and patient couple’s clinical features can significantly improve live birth prediction than using blastocyst images alone. Our next step is to validate the model’s performance via a prospective clinical trial.

## Data availability

All processed data and code needed to reproduce the findings of the study are made openly available in deidentified form. This can be found in https://github.com/robotVisionHang/LiveBirthPrediction_Data_Code, and attached to this manuscript. All codes and software used to analyze the data can also be accessed through the link. Due to data privacy regulations of patient data, raw data cannot be publicly shared. Interested researchers are welcome to contact the corresponding author with a concise project proposal indicating aims of using the data and how they will use the data. The project proposal will be firstly assessed by Prof. Yu Sun, Prof. Ge Lin, and then by the Ethics Committee of the Reproductive and Genetic Hospital of CITIC-Xiangya. There are no restrictions on who can access the data.

## Additional files

1. Supplemenrary file 1. Supplementary Table I and supplementary method.
2. Figure 2-source data 1. Code and data used to generate the ROC curves shown in Figure 2.
3. Figure 3-source data 1. Source data to generate the AUC ranking chart shown in Figure 3.
4. Figure 4-source data 1. Code and data used to generate the heatmaps shown in Figure 4.
5. Figure 4-supplement 1. Two-channel blastocyst images, one channel focusing on the ICM, the other channel focusing on the TE of a same blastocyst.

## Supplementary file 1

## Supplemental method

### Sequential forward feature selection

Fifty-one clinical features were identified to have statistical significance between blastocysts with positive and negative live birth outcomes. The sequential forward feature selection method was used to select a subset of clinical features that are most relevant to the live birth prediction from the 51 clinical features. In the sequential forward feature selection method, features are sequentially added to an empty candidate set until the addition of further features does not increase the prediction accuracy.

We used MLXTEND (http://rasbt.github.io/mlxtend/) to perform the sequential forward feature selection, technical details of the method can be found in Solorio-Fernández et al., 2020.

---

**Figure supplement 1.**
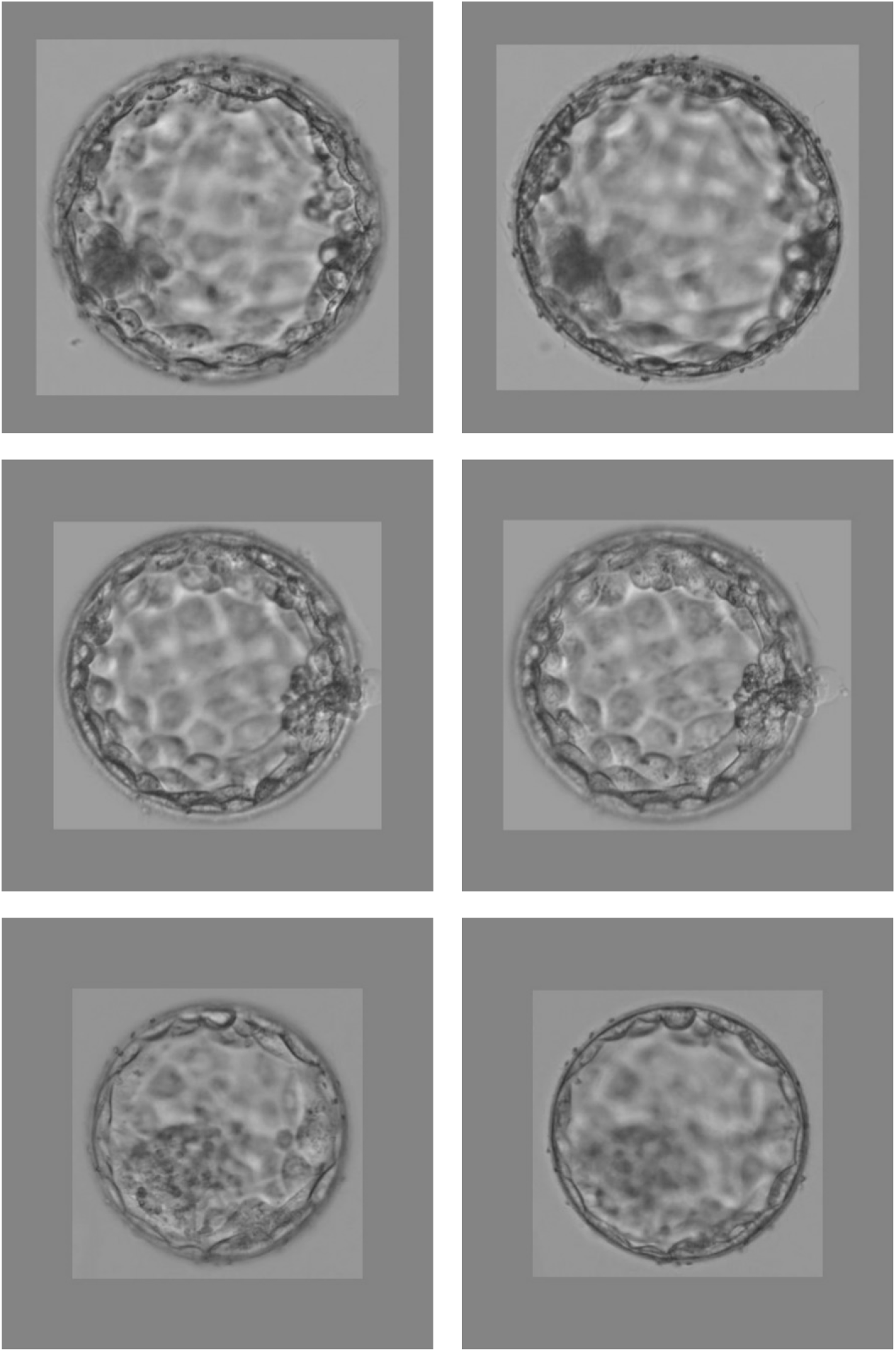
Two-channel blastocyst images, one channel focusing on the ICM, the other channel focusing on the TE of a same blastocyst.

